# The Lived Experience of Implementing Infection Control Measures in Care Homes during two waves of the COVID-19 Pandemic. A mixed-methods study

**DOI:** 10.1101/2021.07.08.21260181

**Authors:** Diane Bunn, Julii Brainard, Kathleen Lane, Charlotte Salter, Iain Lake

## Abstract

**Context:** During COVID-19 care-homes had to implement strict Infection Control Measures (ICMs), impacting on care and staff morale.

**Objectives:** To explore the lived experiences of care-home staff in implementing ICMs.

**Methods:** Mixed-methods study comprising 238 online survey responses and 15 in-depth interviews with care-home staff, November 2020-January 2021 in England.

**Results:** Three themes were identified: ‘Integrating COVID-19 ICMs with caring’, ‘Conveying knowledge and information’, ‘Professional and personal impacts of care-work during the pandemic’. Reported adherence to ICMs was high but fatalistic attitudes towards COVID-19 infection were present. Challenges of providing care using personal protective equipment (PPE), especially for residents with dementia, were highlighted. Interviewees reported dilemmas between strictly implementing ICMs and conflicts with providing best care to residents and preserving personal space. Nine months into COVID-19, official guidance was reported as confusing, constantly changing and poorly suited to care-homes. Care-home staff appreciated opportunities to work with other care-homes and experts to interpret and implement guidance. ICM training was undertaken using multiple techniques but with little evaluation of these or how to sustain behaviour change.

**Limitations:** Results may not be generalizable to other countries.

**Implications:** COVID-19 has had a profound effect on well-being of care-home staff. Despite challenges, participants reported broadly good morale, potentially a consequence of supportive colleagues and management. Nevertheless, clear, concise and care-home focussed ICM guidance is still needed. This should include evidence-based assessments on implementing and sustaining adherence. Groups of care-home staff and ICM experts working together to co-create, interpret and implement guidance were viewed positively.

## Introduction

In many countries the SARS-CoV-2 virus spread rapidly within and between care-homes (CHs) during 2020-2021, causing disproportionate morbidity and multiple deaths amongst staff and residents (Comas-Herrera et al. 2021). UK government support initially focussed on the National Health Service (NHS), ensuring that it would not be overwhelmed by hospitalisations. Initially CHs did not receive the same level of support or preparedness, despite their populations being amongst the most vulnerable to COVID-19 infection (Rajan et al. 2020). Support differed between the NHS and the care-home sector in terms of guidance around infection control measures (ICMs), availability of personal protective equipment (PPE) provision and funding. By the end of June 2020, 31% of all UK COVID-related deaths were in CHs and care-home residents have continued to experience high mortality (Stevenson 2020). High numbers of excess deaths amongst care-home residents have been reported in many countries during the COVID-19 pandemic, likely, if not formally, attributable to the SARS-CoV-2 virus. In international comparisons, deaths in CHs are highly correlated to the total number of COVID-19 deaths in the population living outside CHs, despite differences in reporting between countries (Comas-Herrera et al. 2021; Sepulveda et al. 2020). Contributing modifiable factors in all countries are similar: delays in government guidance and limited specialist support, lack of understanding and misunderstandings around transmission and novel ICM burdens falling heavily on care-home staff (LTC Responses to COVID-19 2021).

Care-home staff have been at the forefront of the pandemic, facing multiple stressors associated with limiting infection transmission and providing care for large numbers of residents being ill and dying with COVID-19, a novel disease where little was known initially and guidance was delayed and continually subject to updates. Care-home staff had to acquire new skills and change working practices to adapt to the demands required of them in a constantly changing environment and with varying levels of support from external agencies (Marshall et al. 2021). Simultaneously, care staff have a responsibility to protect the social environment within residences, because CHs provide the dual purpose of being a home and a centre providing 24-hour care. Implementing, managing and coping with ICMs has posed challenges for care-home staff across all levels and roles, while impacting resident care and staff wellbeing. Staff are key to resident wellbeing and successful implementation of ICMs (Greenberg 2020; Nyashanu et al. 2020). Yet very little research has explored the challenges experienced by care-home staff in implementing and coping with new ICMs during the COVID-19 pandemic. Such information is critical to deal with the current pandemic and enhance preparedness for future events.

Our mixed-methods study aimed to identify the lived experiences of care-home staff (all roles) implementing new ICMs in CHs (including PPE and environmental context), the sources of support available to staff, the effects on staff and those they cared for.

## Methods

This study was approved by our Faculty of Medicine and Health Sciences Research Ethics Committee (reference: 2020/21-038) and registered on the Long-Term Care Responses to Covid Website (10/12/2020; and is reported in-line with COREQ guidelines (Tong et al. 2012). This mixed-methods study comprised an online survey and qualitative interviews with care-home staff, conducted in England during November 2020-January 2021. Both arms were conducted in tandem, data analysed separately and combined subsequently (convergent parallel design). Survey questions and interview topic guide included questions about the challenges encountered in implementing ICMs in CHs, the source of support available to staff and the effects on staff and those they cared for.

The online survey was undertaken using Google Forms, optimised for completion on computers, tablets and/or mobile phones. Recruitment was via posts on our own university social media (Twitter and Facebook), professional and practitioner email distribution lists and eNewsletters targeted at care-home workers across the UK. Respondents were asked to confirm eligibility (aged >18 years and that they worked at least 8 shifts in a UK care-home within the previous four months). We asked respondents to focus on their most recent experiences when answering questions. Survey questions were informed by the Queen’s Nursing Institute survey (Queen’s Nursing Institute 2020), our own previous research (Brainard et al. 2020; E. Smith et al. 2020) and following feedback from current care-home staff (Patient and Public Involvement, PPI).

Survey questions are available on request via corresponding author. We asked about their:

- demographic data (gender, age-band);
- employment data (UK region of workplace, job role(s), years of experience working in CHs, frequency per week of recent shifts, number of CHs worked in during the previous four months; any additional employment where personal care may be required);
- challenges encountered when using PPE;
- morale and emotions;

A validation question was included, asking respondents to click on a certain answer to ensure respondents fully read the questions. Participants were excluded if this question was answered inappropriately. Participation was anonymous, but there was an optional prize draw to win one of five £10 Amazon vouchers. If respondents were interested in taking part in the key-informant interviews, they were asked to provide contact details. We aimed for a sample size of 200. Survey data were analysed descriptively. We explored sub-groups if at least 10 respondents in each category (to ensure anonymity). This allowed us to explore age-band, gender and/or occupation role relationships with PPE use and morale.

For the key informant interviews, we used purposive sampling to ensure we included interviewees in a range of occupational roles and care-home types. Interviews were undertaken using online platforms (Zoom or Microsoft Teams) by two authors (KL, CS). An iterative Topic Guide, developed by the research team, guided the interview and explored:

- how well staff felt prepared to manage and cope with ICMs at the start of the pandemic
- barriers and facilitators to implementing and using ICMs
- experience of working during a possible or actual outbreak (if applicable)
- changes in work patterns and changes in lifestyle
- availability and nature of support

Interviews were recorded, transcribed using otter.ai software, then subsequently checked for accuracy and identifying details removed. All interview participants were offered a £20 Amazon voucher.

After initial interviews, emergent findings and issues were identified together with developments nationally so that key issues could be included in subsequent interviews. This inductive-deductive approach was important because interviews were being conducted during a rapidly changing situation within the UK, including the peak of the second COVID-19 wave and the withdrawal and re-introduction of stringent social-distancing measures.

Interview data were analysed thematically, following Braun and Clarke’s framework (Braun & Clarke 2006) using an inductive approach. All transcripts were coded line-by-line by two members of the research team, DB reviewed the initial set of codes and refined where necessary. Codes were grouped into categories, then sub-themes and themes identified and refined by team discussions. Survey and interview findings were integrated using side-by-side comparisons to provide overall conclusions.

## Findings

The survey and interview studies were not mutually exclusive. We present demographic and background data for the two studies separately, then report joint findings from both study arms under distinct themes identified in the qualitative study. The timing of data collection provides the context for the reported findings. Most data collection occurred near the peak of the second wave: 31% of survey responders and four interviews were completed Nov/Dec 2020; 69% of survey responses and 11 interviews completed January 2021.

### Survey respondents’ demographic and employment details

The survey received 238 unique and valid responses. Although we advertised this survey throughout the UK, 236 respondents resided across England (101 [43%], southern England; 97 [41%] Midlands and East Anglia; 38 [16%] northern England). Non-England residents were retained in the dataset, but the dataset is most accurately described as a survey of care-home staff in England. Most respondents (199, 83.5%) were female, 37 (15.5%) male, and 2 (0.5%) declined to state gender. Table 1 shows the distribution of age-bands reported.

**Table 1.** Age-bands of survey respondents.

| **Age-band** | **Count** | **% of total 238** |
| --- | --- | --- |
| 18-25 years | 44 | 18.5 |
| 26-35 years | 22 | 9.2 |
| 36-45 years | 39 | 16.4 |
| 46-55 years | 62 | 26.1 |
| 56-65 years | 62 | 26.1 |
| age 65+ | 9 | 3.8 |

The majority (215, 90%) had worked in CHs for at least one year, with 177 (74.5%) reporting just one CH job role in the previous four months. Stated job roles were reduced to three broad categories: anyone who ever worked as a senior care-worker or manager (SCW/M, n=164), care-workers who had never worked as seniors/managers (junior care-workers, JCW, n=53), and ‘other’ (any other role, such as administrative, domestic, gardening and health professionals directly employed by CHs, but never worked as care-workers or managers, n=21).

### Interviewees’ demographic and employment details

Recruitment for the interview study was predominantly via the survey, with 62 expressing interest, and a further three making direct contact with the research team. Thirty-six people fulfilling sampling criteria were contacted, and fifteen interviews were conducted (10 female, 4 male, one undisclosed). Interviewees reflected all ages, although all JCWs were under 35 years. As with the survey, interviewees were geographically spread across England. Three had worked in CHs for under a year, and the remainder had varying lengths of experience (2-40 years). Occupational roles included seven in management and eight in frontline caring or domestic roles. Interviewees’ CHs represented a range of sizes, ownership and care provision. Thirteen disclosed that they had experienced a COVID-19 outbreak in their care-home.

### Presentation of themes and integration of survey and interview data

The themes identified from the qualitative interviews are: ‘Integrating COVID-19 ICMs with caring’, ‘Conveying knowledge and information’ and ‘Professional and personal impacts of care work during the pandemic’. Themes were derived inductively, the process involving line-by-line coding, category-grouping, analysis of sub-themes, and finally higher-order themes (Table 2). Themes are presented individually for clarity, and aligned with findings from the survey. Core findings are discussed below. Results from survey questions about challenges encountered when using PPE, staff morale and emotions felt at work were broken down by age-band, gender and/or the three occupational categories.

**Table 2:** Coding and analytical framework for thematic analysis of interview data.

| Themes | Sub-themes | Categories |
| --- | --- | --- |
| Integrating COVID-19 ICMs with caring | Challenges of caring whilst observing ICMs and wearing PPE | personal discomfort of PPE<br>increased time to care<br>effects on residents<br>personal cleaning and hand sanitising |
|  | Dilemmas of caring whilst observing ICMs and wearing PPE | importance of ICMs despite everything<br>prioritising care needs<br>prioritising work-life over home-life |
|  | Impact of social and physical environment | interface between outside world and CHs<br>isolating and cohorting residents within CHs<br>cohorting of staff<br>implementing social distancing<br>cleaning of CH environment<br>location of PPE within CH |
| Conveying knowledge and information<br>(*alternative: “communicating to and within CHs”*) | communicating guidance to CHs | delays in guideline provision initially<br>conflicting guidelines and advice<br>constant changes and updates<br>guidelines lengthy and not care-home-specific<br>external support variable, but named person & manager’s networks most useful<br>COVID-19 champions |
|  | Communicating guidance within CHs | adapting guidelines for specific care-home context<br>adopting a range of communication methods<br>management responsibility<br>ensuring compliance and adherence<br>peer support<br>lack of understanding amongst staff |
| Professional and personal impacts of care work during the pandemic | Coping with COVID-19 outbreak within the CH | speed with which pandemic progressed<br>anxieties directly relating to COVID-19: catching it, being a carrier, protecting own family and protecting residents, impact of ICMs and outbreak within the care-home and on residents<br>financial sources of worry<br>staff shortages<br>undervaluing of care-home staff by society |
|  | External support | family and social networks<br>external agencies |
|  | Supporting each other | financial and other practical support from care-home<br>‘team spirit’ within CHs<br>own resilience |

### Integrating COVID-19 ICMs with caring

#### Challenges of caring whilst observing ICMs and wearing PPE

ICMs and their impact on caring dominated respondents’ and interviewees’ minds. This is unsurprising given the timing of data collection and that reported use of PPE in the survey was high (e.g. 219, 92%, always used masks when working with/near care-home residents, Table 3).

**Table 3:** Reported frequency of use of PPE while working with or near care-home residents, n (%)

|  | Not at all | Sometimes | Often | Always |
| --- | --- | --- | --- | --- |
| Apron | 10 (4.2) | 44 (18.5) | 74 (31.1) | 110 (46.2) |
| Gloves | 3 (1.3) | 29 (12.2) | 63 (26.5) | 143 (60.1) |
| Goggles or face shield | 64 (26.9) | 98 (41.2) | 40 (16.8) | 36 (15.1) |
| Mask | 4 (1.7) | 4 (1.7) | 11 (4.6) | 219 (92) |
| Hand sanitiser | 2 (0.8) | 4 (1.7) | 36 (15.1) | 196 (82.3) |

Challenges were reported around using PPE: 217 (91%) of survey respondents strongly or somewhat agreed that wearing PPE made it harder to give physical care to residents and to communicate with residents or colleagues (Figure 1). Facial PPE (masks/goggles or face shields) was especially linked to difficulties (Table 4).

**Figure 1:**
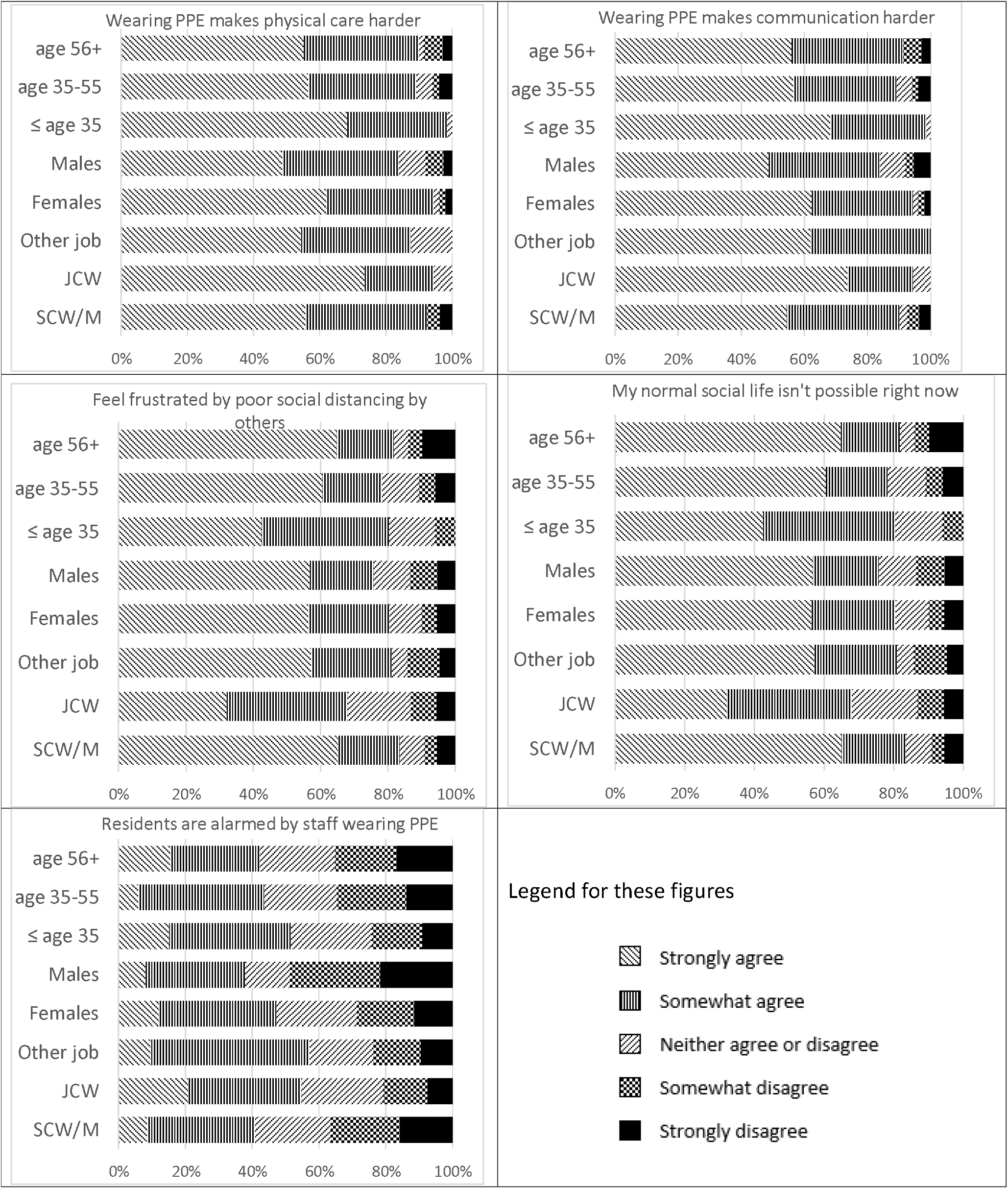
Challenges in wearing PPE and adhering to ICMs. Notes: SCW/M = Senior care-worker or manager; JCW = care-worker who never worked as senior or manager; Other job: care-home staff who never worked as manager or care-worker.

**Table 4:** Reported frequency of difficulties of using PPE when caring, n (%)

|  | Not at all | Sometimes | Often | Always |
| --- | --- | --- | --- | --- |
| Apron | 181 (76.0) | 41 (17.2) | 13 (5.5) | 3 (1.3) |
| Gloves | 164 (68.9) | 54 (22.7) | 15 (6.3) | 5 (2.1) |
| Goggles or face shield | 81 (34.0) | 88 (37.0) | 44 (18.5) | 25 (10.5) |
| Mask | 51 (21.4) | 88 (37.0) | 65 (27.3) | 34 (14.3) |
| Hand sanitiser | 198 (83.2) | 27 (11.3) | 8 (3.4) | 5 (2.1) |

These challenges were highlighted in interviews, with many interviewees reporting unease regarding the effects of PPE on the care they provided.

> *“*..*I do feeding […], prior, I would only wear an apron, […] now I wear mask, apron and gloves, and I have to change all of that every single resident I feed and I feed about 10. So every resident I must take off the gloves, wash my hands, apron, mask, and then reapply them all*.*”* (JCW, i02)

Interviewees found communicating with residents whilst wearing masks difficult, particularly for residents with hearing difficulties and those living with dementia, where verbal communication is aided by facial expressions that can provide additional reassurance and contribute to a sense of safety for residents. Survey responses indicated that many staff thought that residents were alarmed by staff wearing PPE (although fewer SCWs reported this than JCWs or ‘other’ professional groups; 40%, 55% and 57% respectively; Figure 1). One example of this alarm was described by an interviewee who reported that his PPE was torn off him by a resident with dementia, increasing both his anxiety about exposure to infection:

> *“I have had my masks ripped off my visor taken off […], apron, everything, shredded. People are angry, and it just spreads the infection at that point*.*”* (JCW, i01)

Other interviewees described many residents who were initially disturbed by staff wearing full PPE, becoming accustomed to it.

Adherence to ICMs was seen as important in mitigating risk of transmission, engendering feelings of safety. Staff were unhappy about improper ICMs by colleagues or residents. As one JCW stated, *“A lot of people overlook hand-washing and stuff like that*.*”* (JCW, i10)

Time demands needed to don PPE and observe ICMs was frequently discussed. Another key issue mentioned in the interviews was the dilemma between safest practice in terms of ICMs and the interviewees’ caring responsibilities towards residents. Two JCWs described dilemmas between providing safe care if a resident fell with their caring responsibilities. They came to divergent conclusions, with one indicating that donning PPE prior to helping was best practice, whilst the other concluded the reverse.

> *“You can’t go straight up to them anymore like you used to be able to…. Having to go get the PPE and then go back, so you’re effectively leaving people, which is a challenge, […], because it’s not in our nature to leave people*.*”* (JCW, i03)
>
> *“If you see someone falling or fallen, you can’t go, “Hold on a minute, let me get my PPE on” and then help them […]. So it is literally you just have to help them*.*” (*JCW, i08)

The second view was supported by a manager:

> *“We can have a resident who was in distress, and staff are walking away, to go wash their hands, put gloves on, to go back and support that resident […] I don’t care which legislation you’ve got, the resident comes first. If they’re in distress, you support them. Once you’re able to hand them over to another member of staff, you then go and wash your hands*.*”* (SCW, i11)

Other dilemmas between safest practice in terms of ICMs and the interviewees’ caring responsibilities towards residents emerged when caring for residents nearing the end of their lives. One participant describing how they abandoned full PPE in these situations even though the resident had COVID-19.

> *“I hold my hands up when we have people dying. […] I wasn’t remotely surprised I got it. […] But I just, I guess I thought it’s just part of what we do. I wanted to give someone compassionate care […] at the end of their life*.*”* (SCW, i12)

Conflicting priorities and knowing when and how to adapt ICMs to provide safe care was a source of anxiety. Being aware of this, this same manager described how she included rationales in her staff training to empower staff to make informed care decisions in such situations. She emphasised the importance of *‘Explaining to them the reasons behind what we’re doing, when we’re doing it and how we’re doing it*.*”* (SCW, i11)

#### Impact of social and physical environment

Adaptions to care-home life also presented dilemmas to staff. Many CHs ‘de-cluttered’, worried that active virus remaining on objects would facilitate transmission. For some interviewees, this was a step too far – changing the environment from a home into a mini-hospital:

> *“Then we removed anything that […] residents could touch to stop the spread. We are still trying to make it feel as if it’s their home, because that was the most saddest part, that was the most cruel thing, was removing the comfort things that they had, excuse me, makes me upset*.*”* (JCW, i14)

In interviews, cohorting and maintaining social distancing produced mixed feelings and interpretations. Cohorting in the main was not viewed favourably, as it was seen as challenging in older buildings that did not permit ready alteration, and an intrusion for residents. It meant moving residents from their home (own rooms) and was particularly disruptive for people living with dementia, causing further confusion and disorientation. One SCW (i04) said her *“…. old vicarage-type building […] doesn’t lend itself to isolating people at all*.*”* Another (JCW, i03) stated that cohorting

> *“…*..*was never going to work, because people have their own rooms, and that’s their home, isn’t it? […] They’ve got all their things in there. […] So yeah, that plan kind of came and went*.*”*

Social distancing was considered important for residents, but it meant some residents had to be isolated in their rooms at certain points in the pandemic, especially when moving into a care-home or after any hospital visits (either outpatient or inpatient). Views on the importance of social distancing between staff were varied; on the one hand, changes were made within CHs to ensure this (moving meeting rooms, discouraging staff from car-sharing or dining with residents), but other interviewees described how social distancing between staff was not possible due to prioritising different concerns.

> *“The biggest thing is that we don’t socially distance. […] but there’s nowhere else in the building to do handover. It’s confidential, you have to do that*.*”* (JCW, i05)

When asked about social distancing in the wider community, 190 (80%) of survey respondents reported feeling frustrated by others’ poor social distancing practices (Figure 1).

Managing the interface between the outside world and the care-home to minimise the risk of introducing COVID-19 into the care-home featured strongly in all interviews, with interviewees describing many extra precautions they and their families took to keep residents, colleagues, themselves and their own families safe. For many this was seen as a duty:

> *“I took the rules outside of the building very seriously… I didn’t socialise, and I felt a duty to do that due to our work. And you know, my family also did that, because of where I work*.*”* (JCW, i02)

In the survey, 188 (79%) reported that they strongly or somewhat agreed that their social life was affected by the pandemic, although amongst JCWs, this was lower (68% compared to 83% and 81% amongst SCWs and ‘other roles’, respectively; Figure 1).

Prevalence of cross-setting employment was addressed in the survey, with eighteen (7%) respondents reported working in >1 care-home in the four months prior to completing the survey, and 41 (17%) reported working in other care settings (eg., NHS, domiciliary care, childcare).

Reduced or restricted visiting CHs were widespread during the pandemic and extended to health professionals (HP), as well as families, volunteers and maintenance people. Fewer visits were viewed positively in several instances, particularly HP visits, as HPs were seen as high-risk carriers (visiting multiple homes) where *‘…*.*those sort of jobs [i*.*e*., *HP visits] just mean that there’s more people coming in, which obviously is a bit of a risk’*. (JCW, i06)

Fewer visits freed up care staff time as managing visitors to the home was a source of anxiety requiring extra facilities, added expenditure, staff training, and extra staff time in managing the whole process.

### Conveying knowledge and information

The aim of knowledge exchange is to inform and change practice for the better. Conveying knowledge and information during the rapidly changing landscape of the pandemic has caused widespread difficulties everywhere, not least the care-home sector, as official guidelines are constantly changed and updated.

#### Communicating guidance to CHs

A major factor for all interviewees, particularly those in management, was the need to adapt official guidance for their own particular care-home. They said that generic guidelines simply could not otherwise be implemented, due to being too broad, not written with the care-home sector in mind, and therefore inappropriate. Adapting them was resource-intensive, especially during staff shortages, further draining the physical and mental reserves of those in care-home leadership roles. Many SCWs were frustrated that policy-makers had not appeared to consult or work with care-home staff when creating guidance, with one manager stating: *“I don’t think there was enough opportunity to ask the people doing the job to voice what they thought would help*.*”* (SCW, i14). Another reported:

> *“You’d spend three or four hours going through stuff and then pick out a couple of paragraphs that are applicable to you. The rest isn’t. So I just wasted all that time. I don’t really have much time anyway*.*”* (SCW, i13)

Additional anxieties discussed were the ‘piecemeal’ approach to guidance provision, multiple sources of guidance from various authoritative bodies and perceived conflicts between them, continuous updates, delays in care-home-specific guidance and differences in implementation between CHs (even those within the same company), as well as lack of trust in media reporting. All this caused confusion, and engendered apprehension amongst staff of missing something crucial.

> *“Because the guidance kept changing, do I think that it was muddled or made us feel muddled? Yes, I do. Because we were asking ourselves, “is this right?” […] Some of those guidelines that were introduced, say at 12 o’clock, by one o’clock, “do not follow those guidelines”. But you’d already implemented those with your team! Then they’re like, “This is an update”. So, you know, this is why I think the information should have been on a daily bulletin*.*”* (JCW, i14)

Confusion extended to critical areas such as whether staff could be reimbursed for self-isolating, a key ICM for preventing COVID-19 spread. Funding has been provided for staff self-isolating, but as one manager pointed out, *“The understanding of the use of the infection control funds is very, very disjointed, in my opinion”*. (SCW, i04)

Many SCWs discussed strategies they had created or accessed to support them in adapting and developing their own care-home-specific guidance. Managers relied on networks of managers (informally or formally, if part of a larger company), with one manager explaining how her company *“…picks out the pieces that we need*.*”* (SCW, i15). A number of managers placed particular value on being able to contact a named person, whose advice could be trusted and relied on, seen as *“…a great font of knowledge [who] we always go to*.*”* (SCW, i15). In one area, this was a local respiratory consultant, in another, it was the local authority infection lead.

#### Communicating guidance within CHs

Cascading information down to all staff was seen as a care-home management responsibility, together with ensuring that staff adhere to best practice: *“You’ve got to do everything you can to make it safe*.*”* (SCW, i15). JCWs were aware of the need for guidance to be adapted, relying on their senior colleagues to provide appropriate guidance for their care-home.

> *“Yeah, we got all the information we need and they [senior staff] told us how we need to act, and how I need to work now and the changes that have been made*.*”* (JCW, i02)

This was mirrored in the survey, where 84% felt that their managers were supportive in preventing COVID-19 (Figure 2). Interviewees described various ways in which guidance and ongoing changes were communicated within the care-home including formal training and shadowing more experienced staff. However, it was acknowledged that the quality of the latter type of training was variable.

**Figure 2:**
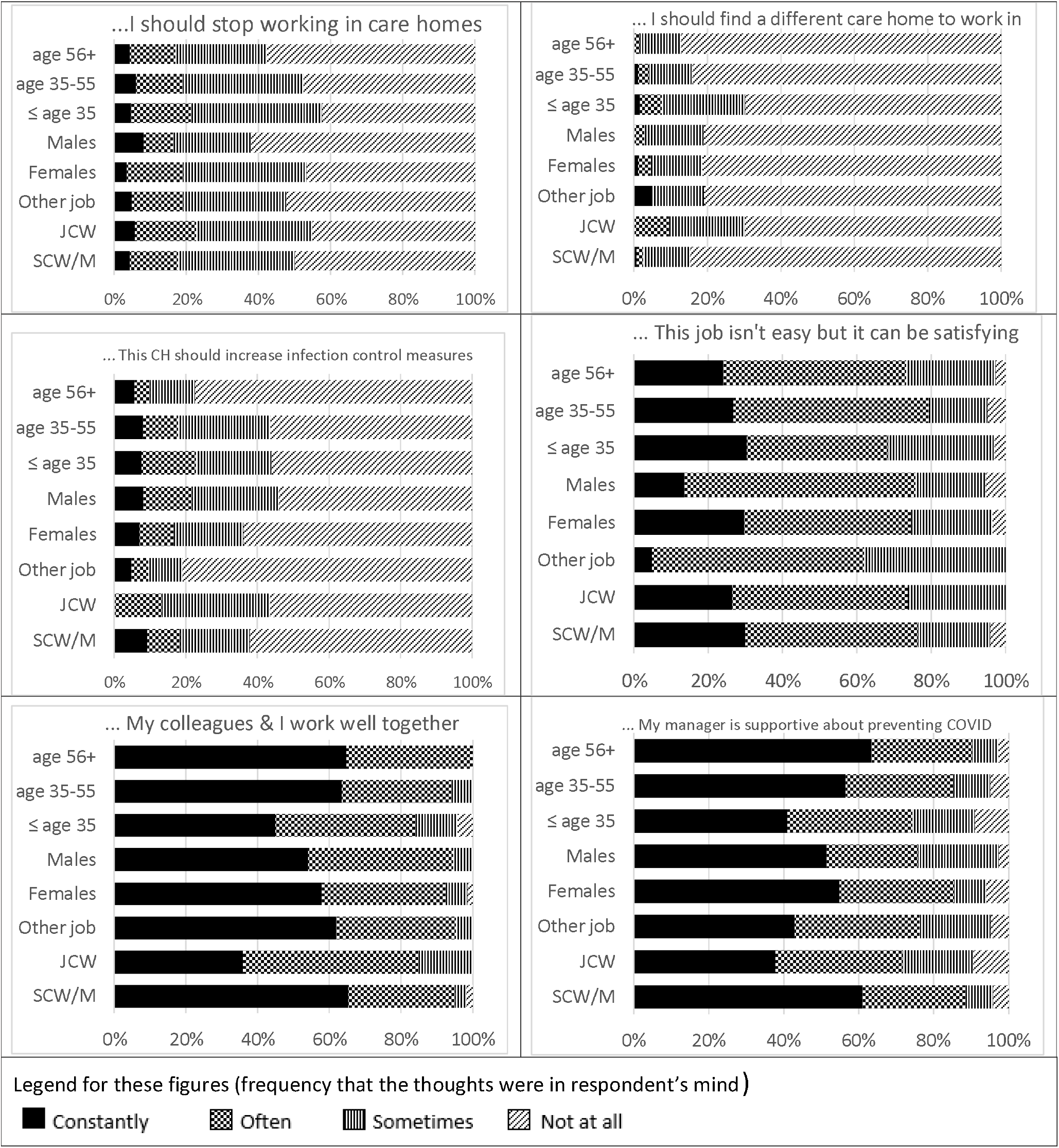
Thoughts on morale and attitudes respondents held recently about work. Notes: SCW/M = Senior care-worker or manager; JCW = care-worker who never worked as senior or manager; Other job: care-home staff who never worked as manager or care-worker. “Work well together” refers specifically to working together to prevent COVID-19.

> *“I wasn’t actually sat down and told how to do it. Like I said, I was lucky enough to have someone who I was shadowing who was very helpful and helped me but if you had somebody who you were shadowing who just sort of went, here you go, […] it could be difficult*.*”* (JCW, i08)

Three JCWs felt they were responsible for updating themselves, including one who saw their manager as too complacent and so not trusted to make the home as safe as it could be. This JCW then described how they tried to suggest some changes and were frustrated by the limits of the information they received:

> *“I’d talk to management: is this something we should be doing/could we be doing this? I have a great deal of information from previous training I’ve done, work experience in different homes. […] The only information they [management] made available to us was about what was legal (having to wear masks in the car; not allowing visitors)*.*”* (JCW, i08)

On the other hand, some managers acknowledged their frustrations around producing guidelines which were not read or implemented, although all agreed that time for training was considered paid worktime. This was a moot point when many regularly worked beyond their formally contracted hours.

> *“Initially when I came here, I was putting it all into a big folder, and people were so busy, so nobody was seeing it. And if we were giving them training to do, they weren’t doing it. So what I do now is every time I get an update, I email it directly to their personal emails*.*”* (SCW, i04)

Interviewees discussed the various methods of communications used within their CHs, many developed by trial and error as to what worked best, acknowledging that different approaches were essential as people learn in different ways. Five interviewees described ‘COVID Champion’ roles -key people who could influence, empower and support colleagues with COVID-related activities, such as ICMs and testing. Many interviewees discussed a form of peer-monitoring – ‘keeping an eye’ on each other and supporting their colleagues, acknowledging that lack of compliance likely reflected unawareness or forgetfulness. This reinforced the view of many managers that criticism was unhelpful, and a positive supportive approach was needed to better support staff and ensure compliance and adherence to ICMs.

> *“And we’ve not here taken any disciplinary action against anybody. And we know people, you know, flout it a bit. […] I think we’ve built up quite a repertoire of help and assistance that we give to staff*.*”* (SCW, i04)

Staff at all levels discussed that due to longevity of the pandemic, constant vigilance about ICMs was stressful. Several managers (but no JCWs) described implementing competence and adherence checks, such as hand-washing audits, to ensure ongoing compliance. One manager said this was in response to their insurer’s request. Many interviewees strongly relied on implementable, practical ICM procedures, particularly cleaning activities (hand-washing, sanitiser use, quarantining deliveries). Possibly these measures attracted attention because they are known to be effective in preventing transmission of other infections, such as norovirus, which care staff are familiar with. In spite of continuing deaths in CHs, very few (40, 17%) survey respondents felt that their CH needed to increase their ICMs (Figure 2).

### Professional and personal impacts of care work during the pandemic

Care-home staff found working during the pandemic stressful. Some sources of stress and anxiety are highlighted above (impact of ICMs on care, implementing continuous changes in guidance). Though many others were reported, the most overwhelming experience was dealing with an outbreak.

#### Coping with COVID-19 outbreaks within CHs

Thirteen interviewees had experienced a COVID-19 outbreak in their care-home, ranging in extent and severity. Some interviewees became emotional while describing how they coped with multiple losses of life amidst staff shortages, increased workloads, conflicting guidance and variable support from external agencies. One JCW described losing half their residents over just a few weeks:

> *“We did lose probably about four or five that wasn’t COVID, yeah. But after that, it just took them all. […] It’s been so bad, I can say that, devastating. […] we see them every day, it’s just so sad*. (JCW, i14)

Experiencing multiple deaths is particularly poignant when care-home staff develop long-term relationships with residents. One JCW (i14) encapsulated this, saying *“…these residents are my family too*.*’*

The speed with which an outbreak spread was unforeseen described by one SCW (i11) as *“Very scary”* adding to the emotional toll on staff.

> *“I don’t think anything could have prepared us really for how many people and how rapidly it has spread*.*”* (SCW, i12)
>
> *“We had well over 30 staff and we were down to 7 because we all went off with COVID*.*”* (JCW, i10)

With such major shortages of staff, employing agency staff became inevitable, despite it being well-known that this increased risk of transmission into the care-home:

> *“We have a fantastic staff team who are picking up shifts where there are shortfalls to try and prevent and reduce having to use agency staff, which may be working in other homes. […] We need to stop agency coming into the home wherever possible*.*”* (SCW, i11)

An outbreak created substantial staff shortages due to increased resident care needs and staff sick or self-isolating. This had widespread financial implications – the cost of employing agency staff, financial recompense for permanent staff and increased costs of PPE (purchasing and disposing) and other ICMs. Whilst most staff reported receiving remuneration when off sick or self-isolating, this was not universal amongst the interviewees – mainly JCWs, some of whom were unsure about what recompense they could expect. As discussed previously, this uncertainty likely relates to confusing guidance. This was reflected in the responses to an interview question where we asked for participants’ views on a news report where care-home staff had worked whilst symptomatic for COVID-19 (https://www.bbc.co.uk/news/uk-england-lancashire-55072118). Whilst most condemned the practice, there was also some understanding, particularly from JCWs (typically among the lowest paid) signalling the conflicted position some found themselves in.

> *“I’ve had so many people say to me in my personal life, ‘I’m still going to work even if I test positive because I can’t afford not to work’*.*”* (JCW, i05)

This contrasts with fears and anxieties expressed by many participants about being the person to transmit infection (either from the care-home to families or vice versa). Interviewees and their families took precautions in their personal lives which took a toll on interpersonal relationships. One participant described sending their children to live with grandparents, whilst another moved away from their family.

> *“I had a lot […] of anxieties about being the person to bring it in. […] Every precaution we probably possibly could have taken. I didn’t see any family members. I didn’t see my partner. I didn’t see my sister […] I had no interaction*.*”* (JCW, i02)

For other care-home staff, there were elements of fatalism and inevitability regarding contracting COVID-19, as one JCW (i10) reported: *“Working in a care-home just comes with that risk at the minute*.*”* Another JCW, who contracted COVID-19, was told by a senior colleague:

> *“Oh, well, you all should have expected this was going to happen at some point, you all knew you were going to be working with positive patients*.*”* (JCW, i08)

The cumulative effect and relentlessness of all these stresses (and others), over a year, on staff resilience was reported frequently by both SCWs and JCWs:

> *“I burned out in the end. […]. In hindsight, it was so draining because it was constant. And it was constant fear, and I think fear is the just the worst thing, the fear from getting ill and my actions making someone ill – or my lack of actions, I should say. That’s a horrible thought to have*.*”* (JCW, i05)

#### External support

Care staff looked for support from many external organisations (e.g., Care Quality Commission, local and national government, NHS), which typically was forthcoming and appreciated. Nevertheless, six interviewees described how dealing with conflicting views from health and social care professionals, alongside unhelpful and critical attitudes, created tensions and additional burdens, contributing to their stress and isolation. As one SCW who had experienced a major outbreak explained:

> *“In our area they put together […] an MDT [multi-disciplinary team] to support a home with an outbreak. […] there was a level, not from all parties, […] of criticism towards staff. […] but you also felt that they were looking for any opportunity to find faults. For us, […] I didn’t find it a particularly positive experience*.*”* (SCW, i12)

HPs were not the only source of criticism. Many interviewees felt that care staff received unfair criticism from multiple parties, as one manager pointed out:

> *“So we’re getting it from residents. We’re getting it from families. We’re getting it from visiting professionals*.*”* (SCW, i11)

Anger was expressed by some interviewees, and in the survey, feeling angry (either ‘constantly’ or ‘often’) was reported by 62 (26%) of staff, but in much higher levels by ‘other’ occupational roles compared to SCWs or JCWs (33%, 19% and 15% respectively, Figure 3) and those aged 56+ years (27%, compared to 18% in each of the lower age-bands).

**Figure 3:**
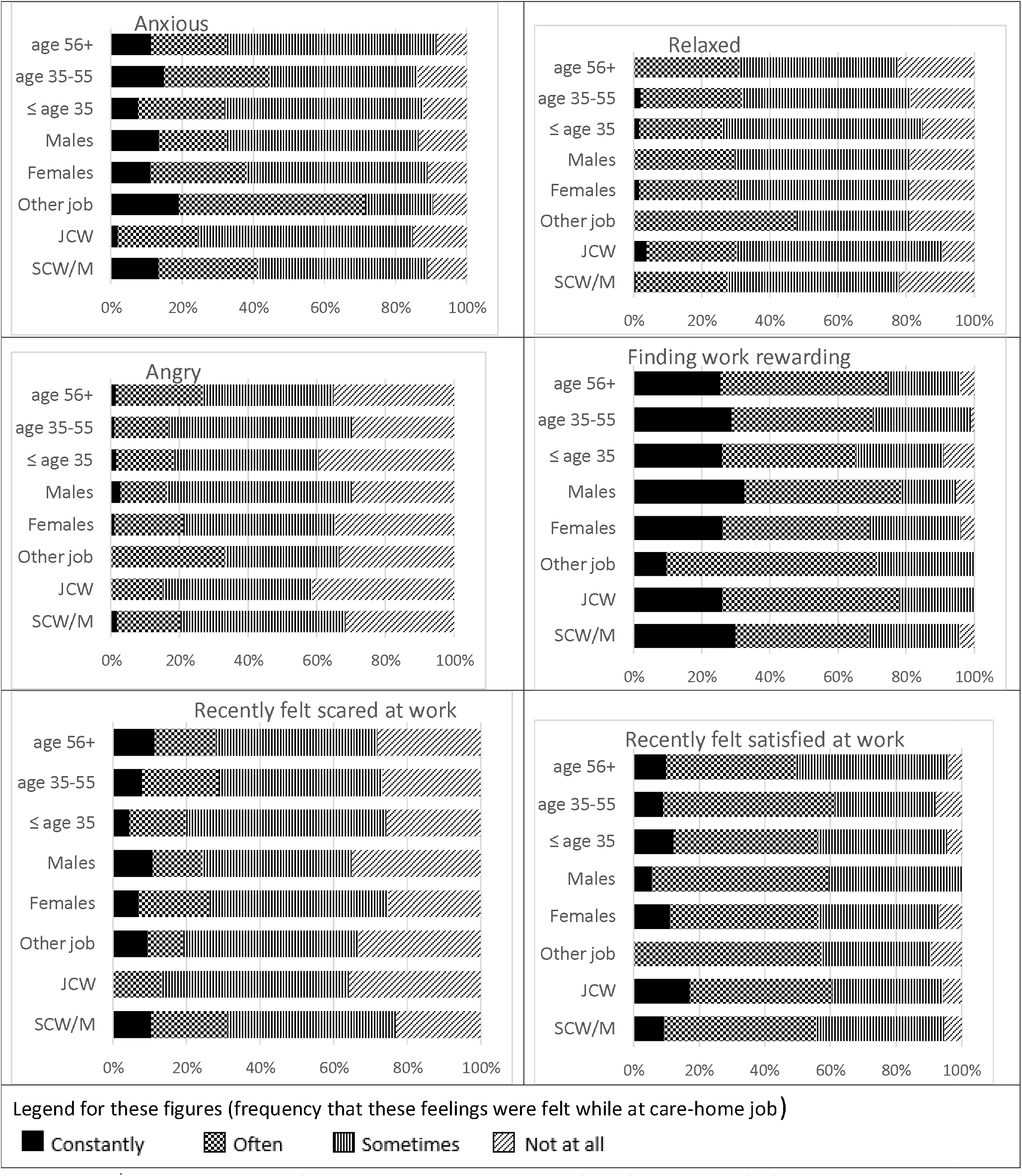
Emotions that respondents felt recently while at work. Notes: SCW/M = Senior care-worker or manager; JCW = care-worker who never worked as senior or manager; Other job: care-home staff who never worked as manager or care-worker.

Nonetheless, interviewees valued the support they received from many quarters, including HPs, families and the wider community. One village was described as *“absolutely brilliant”* (SCW, i04) for offering support, including shopping for CH workers.

#### Supporting each other

Almost all interviewees valued supportive colleagues and managers, indicating a ‘pulling together’ of staff, and team spirit. Many interviewees cited explicit examples of support, from the very practical to more focused attention on well-being. This ranged from one company that *“organised mental health and well-being counselling service”* (SCW, i11) to another that *“provided pasta, eggs, milk, a lot of food groceries for us during COVID*.*”* (JCW, i02). In addition, managers reported placing great importance on a trusting, supportive relationship between themselves and their deputies (we did not interview any deputy managers).

This internal support between colleagues was reflected in the survey, where 93% indicated that they worked well with colleagues (Figure 2). However, colleague support was not universal: two JCWs noted lack of support, expressing concerns about poor practice and feeling unsafe and lack of trust.

> *“A member of staff was immediately sacked for an incident. I don’t think we could trust each other. It definitely, it did have an impact*.*”* (JCW, i05)

The survey asked about many possible emotional states while at work. For SCWs, 31% reported feeling scared and 41% felt anxious (either ‘constantly’ or ‘often’), much higher levels than those reported by JCWs (13% and 25% respectively) although it was more mixed in the ‘other’ occupational roles (19% and 71% respectively). No marked differences surfaced between ages or genders for any of these emotional state questions (Figures 2 and 3), so the differences between SCWs and JCWs likely relate to leadership roles and feelings of greater managerial responsibility in ensuring that staff and residents were safe. One-third of respondents felt relaxed at work. Respondents were also asked about whether they thought about leaving care-home work or moving to a different care-home, with 19% and 4% ‘constantly’ or ‘often’ reporting these thoughts, with no differences across occupational, age or gender groupings. This corresponded to positive emotions about work, with 56% ‘constantly’ or ‘often’ feeling satisfied with work recently, even when 74% acknowledged that ‘the job was not easy, but satisfying’ (Figure 2), and 70% thought their work rewarding. Both older (56+) and younger workers (<35 years) were less satisfied than those aged 36-55 years (Figure 3).

## Discussion

This is one of the first studies to explore the lived experience of care-workers living and working with ICMs. Its purpose is to enhance our understanding of factors impacting on SARS-CoV-2 transmission within and between CHs. Using a mixed methods approach, we triangulated data from 15 in-depth qualitative interviews with 238 survey responses to provide a rich analysis of care in UK CHs during COVID-19. The qualitative interviews provided contextualised experiences and meanings, particularly relevant when deepening our understanding of an international pandemic which has had a profound impact on those living and working in CHs, and essential for policy makers to acknowledge (Tremblay et al. 2021).

Our findings should be placed within the context of the data collection period (December 2020-January 2021) coinciding with the height of the second wave of COVID-19 in England, the third national Coronavirus lockdown starting 06/01/2021, routine testing in place and the vaccine programme commencing 08/12/2020, so disease awareness was especially high amongst participants.

A key strength of this study is the inclusion of many JCWs in both the survey (22%) and interviews (53%). Whilst our recruitment under-represents JCWs (estimated as 76% of the UK social care workforce) (Skills for Care 2020), this is still one of the few studies to have included these care-workers, as difficulties of recruiting JCWs into studies is well-documented (Lam et al. 2018). We have identified where JCWs’ views were particularly pertinent or differed from those of SCWs in our report, although many SCWs were also involved in direct care throughout the pandemic.

The integrated survey and interview findings were presented under three themes: ‘Integrating COVID-19 ICMs with caring’, ‘Conveying knowledge and information’, ‘Professional and personal impacts of care work’ (Table 2). In our first theme, survey responses indicated high adherence (e.g. mask-wearing, 92%, Table 3) when working near CH residents, higher than previously reported in a survey conducted June 2020 with workers working across multiple healthcare settings (i.e. not CH-specific) (L. E. Smith et al. 2020). In our study, fatalism regarding inevitability of contracting COVID-19 was highlighted, and this is problematic as fatalism is associated with lower adherence to ICMs (L. E. Smith et al. 2020), but whether this is cause or effect still needs to established. A fatalistic attitude may limit adherence to ICMs, but a lack of appropriate ICMs or understanding of ICM use may also increase COVID-19 risk, thus reinforcing the inevitability of contracting the disease.

We reported on the challenges around providing care whilst adhering to ICMs and wearing PPE, such as resident distress, staff discomfort and dilemmas around prioritising care. Resident distress was particularly apparent for those living with dementia who were disturbed by social distancing, the visual appearance and impaired communication associated with staff PPE use, although some interviewees reported a degree of accustomization amongst residents. Whilst these findings resonate with those reported by other CH studies (Marshall et al. 2021; Nyashanu et al. 2020), a key feature of several interviews in our study (both JCWs and SCWs) were the daily dilemmas reported by CH staff about when strict implementation of ICMs seemed to negatively impact on care, creating dilemmas around prioritising conflicting care needs. Whilst others have reported on some of these issues (White et al. 2021; Marshall et al. 2021), the everyday care dilemmas and knowing which aspects of care to prioritise, to provide person-centred care for individual residents, were reported on frequently in our study, indicating the complexities of decision-making which CH staff are involved with. This has ongoing impacts when staff feel they may not have made the ‘right’ decision, thus increasing anxiety and stress. Further research in how best to support staff in making these decisions to provide best practice is needed.

Wider dilemmas about how and when the CH’s social and built environment should be adapted to accommodate best practice in ICMs that fit in and encompass the fundamental tenet that CHs are ‘homes’ also need to be addressed, and be at the forefront of CH design going forwards (Anderson et al. 2020). A further critical ICM to prevent the spread of COVID-19 in CHs are payments to ensure that CH staff are paid if they have to self-isolate due to illness or suspected COVID-19, yet the Interview data highlighted confusion around this key measure and how to access the Adult Social Care Infection Control Fund^1^.

The second theme (conveying knowledge and information) found that, nine months into the pandemic, official guidance was still reported as confusing, constantly changing and needing to be interpreted by senior care-home staff. Guidance was perceived to be too general and not attuned to CH settings, particularly the challenges of residents living with dementia. This was reported earlier in the pandemic (Spilsbury et al. 2020; Marshall et al. 2021; Rajan et al. 2020), and remained an issue. Yet, during any crisis effective and clear guidance is essential to staff reassurance and confidence (Bryson 2018), and including representatives from the CH sector in guidance development is essential.

Where possible, staff appreciated the opportunity to work together with other CHs and experts (e.g. infection control specialists) to interpret guidance, problem-solve and share best practice, and this was found to be extremely beneficial. Within CHs, the constantly changing guidance was cascaded down to staff, using a ‘toolkit of methods’ adapted to fit in with availability of in-house resources, and staff with a range of roles and mixed learning abilities. Examples included providing access to online materials, designated COVID champions and shadowing more experienced staff, although varying levels of success were reported for these measures. We recommend that further research is needed to identify effective methods to train and communicate with all staff, not just to impart information but also to empower staff and support their confidence in informed, evidence-based decision-making to enhance care. In turn, this will support mental health and wellbeing (Pollock et al. 2020). Additional research is required to determine how to sustain behaviour change, highlighted as instrumental in addressing the continuous threat of COVID-19 (Michie & West 2020), and identified by many interviewees as an ongoing source of anxiety.

The final theme (professional and personal impacts of care work during the pandemic) reported on how the pandemic had impacted on the lives of care staff. Fear of infection (transmitting and being infected) was frequently reported, as it has been in other studies of frontline healthcare workers (Nyashanu et al. 2020) but the greatest distress came from staff who had experienced a COVID-19 outbreak within their CH. The huge impact this has had on well-being has been recognised, but many more measures need to be in place in both the short- and long-term to support CH staff (Spilsbury et al. 2020; White et al. 2021; Marshall et al. 2021). Some of these measures should address causative factors, such as appropriate guidelines and staff shortages, but other measures are urgently needed to provide ongoing support in addressing moral injury and post-traumatic stress disorder (Pollock et al. 2020). In spite of all these difficulties, our survey respondents reported broadly good morale and more positive than negative emotions about their work (Figures 2,3). Many (but not all) interviewees reported on the positive effects of supportive colleagues, management and external support. This is important as recognition and support are commonly cited as important factors in motivating healthcare workers (Afolabi et al. 2018), and especially during the pandemic (Marshall et al. 2021; White et al. 2021; Spaetgens et al. 2020).

Whilst our study has focused on personal experiences of care workers during the pandemic, wider contextual factors undoubtedly impact on these. Spilsbury et al. (2020) sought to identify and categorise lessons learnt about COVID-19 management, working with NHS and senior CH staff, and concluded that underpinning systemic issues of staff shortages, underfunding and under-valuing of the CH sector were contributory factors, issues that were highlighted by many of our interviewees.

## Conclusions

The COVID-19 pandemic has had an unprecedented effect in CHs. Direct-care staff have faced many challenges in coping with their own and their resident’s exposure to the disease. Transmission of COVID-19 within and between CHs is impacted by the response of care staff to the many ICMs, how these are communicated and how they impact on care. Beyond the CH, a more informed and understanding approach by government to the distinct settings found in social care is needed to support reductions in transmission. Our recommendations are outlined in Box 1.

## Data Availability

Subject to not yet obtained approval by our IRB, the survey data and thematically coded data may be available from authors.

## Acknowledgements

We would like to thank our care workers who piloted the survey and all the survey and interview participants for generously giving their time.

Dr Bunn: this is a summary of research supported by the National Institute for Health Research (NIHR) Applied Research Collaboration East of England in collaboration with the University of East Anglia. The views expressed are those of the author(s) and not necessarily those of the UEA, NHS, the NIHR or the Department of Health and Social Care.

Professor Lake and Dr Brainard were funded by the National Institute for Health Research Health Protection Research Unit (NIHR HPRU) in Emergency Preparedness and Response at King’s College London in partnership with Public Health England (PHE) and collaboration with the University of East Anglia. The views expressed are those of the authors and not necessarily those of the NHS, the NIHR, UEA, the Department of Health or PHE.

## Box 1

**Recommendations for practice**

The constantly changing guidance and longevity of ICMs throughout the pandemic was unprecedented. Findings from this study can inform future strategies. We recommend:

- Clear, concise, care-home-focused guidance, informed directly by staff working on the frontline within the care-home sector.
- Guidance relevant to people living with dementia
- Revised guidance released periodically (e.g. weekly) as opposed to on an ad hoc basis.
- Evidence-based advice on cascading and implementing guidance to staff using a variety of methods.
- Advice on empowering staff to make informed decisions when faced with care dilemmas.
- Clarity about reimbursement for staff undergoing training, self-isolating, sick leave.
- Sustained support networks within and outside care-homes.
- Identify a knowledgeable key point of contact for managers.
- Work to improve and develop positive supporting relationships across health and social care.
- Promote the complexity of care within care-homes, to enhance public and health and social care professionals’ perceptions.
- Support preparedness for future outbreaks.

## Footnotes

1 Established by the government in June 2020 to support providers to reduce rates of COVID-19 transmission, https://www.gov.uk/government/publications/adult-social-care-infection-control-fund.

## Notes

***Conflicts of Interest:***

### Competing Interest Statement

The authors have declared no competing interest.

### Author Declarations

Faculty of Medicine and Health Sciences Research Ethics Committee (reference: 2020/21-038)

